# Mortality & COVID-19: A Snapshot of a Tertiary Care Facility in Pakistan

**DOI:** 10.1101/2020.08.25.20181792

**Authors:** Muhammad Asim Rana, Ahad Qayyum, Mubashar Hashmi, Muhammad Muneeb Ullah Saif, Muhammad Faisal Munir, Rizwan Pervaiz, Muhammad Mansoor Hafeez

## Abstract

**Introduction:** Ever since Sars CoV-2 infection has started from China and has taken the shape of pandemic the mortality associated with this disease has been under discussion and hypercoagubility, acute severe respiratory syndrome and sepsis with multi organ failure have been accursed as possible reasons of deaths in cases infected with novel Corona virus. We conducted a retrospective analysis of the cases admitted in our high dependency and Intensive care unit and tried to pinpoint the major cause of mortality in our cases.

**Methods:** This is a single center retrospective study carried out at Bahria International Hospital Lahore over a 3 month period (May 10^th^ to July 10^th^ 2020) in which we analyzed the clinical and biochemical profiles of the COVID-19 patients who died during this period.

**Results:** A total of 108 patients were admitted during this period out of which 11 patients died. 7 of them were men and 4 women. Majority of them had sudden cardiac arrest due to acute coronary syndrome followed by multiorgan dysfunction syndrome and acute respiratory distress syndrome.

**Conclusion:** Acute coronary syndrome due to hypercoagubility was the leading cause of death in our patients.

## Introduction

Coronavirus diseases 2019 (COVID-19), also known as Severe Acute Respiratory Syndrome Corona virus 2 (SARS-CoV-2), is the reason for the global pandemic that started from Wuhan in December 2019. Despite the fact that the majority of patients undergo an uneventful recovery, in around 19% it leaded to severe pneumonia and in 14% there is a progressive worsening critical pneumonia(1)

To date more than 20 million cases of COVID-19 have been confirmed globally with the fatality number touching one million deaths. Various specialists have detailed diverse case casualty proportion ranging from 4.2% to 11%(2). A national level review in china which included 1099 patients with COVID - 19 has referenced that 5% of cases were sufficiently serious to be admitted to ICU and their casualty rate was 1.36% (3) The total number of COVID19 patients to date are approximately 0.3 million with under 7000 deaths in total(4).

The virus belongs to identical genus as severe acute respiratory syndrome coronavirus (SARS-CoV) and Middle East respiratory syndrome (MERS)-CoV, and was thus named SARS-CoV-2 by the International Committee on Taxonomy of Viruses on February 11, 2020. However, SARS-CoV-2 is more infectious than SARS-CoV and MERS-CoV, with over 84,000 cases in China and over 4,200,000 cases worldwide reported as of May 12, 2020, per the middle for Systems Science and Engineering at John Hopkins University(5). On February 11, 2020, the World Health Organization officially named the disease caused by the new coronavirus as coronavirus disease (COVID-19). SARS-CoV-2 is prone to transmit in family clusters or cause outbreaks in hospitals(6),(7). Most patients with COVID-19 present with mild and moderate symptoms, but severe cases can present with acute respiratory distress syndrome (ARDS), multiple organ dysfunction syndrome, and even death. it’s been reported that death rate of COVID-19 varies from 1.4% to 4.3% in several regions or hospitals(8).

## Methodology

A retrospective study conducted in Bahria International Hospital Lahore. The study was approved by the ethical committees of hospitals. All consecutive patients with severe, confirmed COVID-19 admitted to hospital from 10^th^ May till 10^th^ of July 2020. COVID-19 was confirmed through nasopharyngeal and throat swab samples by real time RT-PCR (9). In the present paper we aim to review and highlight the clinical and biochemical characteristics of our fatal cases. Bahria International Hospital is a major trust hospital in the second largest city of Pakistan i.e. Lahore. We dedicated 20 high dependency unit beds and a 10-bedded ICU for the care of the Corona patients. Clinical and laboratory test results, as well as clinical management data, were obtained using data collection forms from medical records. The data were reviewed by physicians and Bahria International Hospital. Data collected included demographics, medical history, underlying comorbidities, laboratory findings, and clinical management. Continuous measurements, such as mean (SD) and categorical variables, were reported as numbers and percentages (%). For laboratory results, we also assessed whether or not measurements fell within the normal range. We used SPSS (version 21.0) for all analyses.

## Results

We admitted 108 number of cases in Bahria International Hospital Lahore, Pakistan from tenth May to tenth of July. 30 were moved to ICU and out of these 11 surrendered to death because of pandemic disease. In the present paper we aimed to review and explore the clinical biochemical characteristics of our fatal cases. Bahria International Hospital is a small corporate hospital in Lahore. We created 20 beds isolation rooms and 10 bed ICU to help and treat patients infected with Corona Virus. 11 fatal cases were included. The mean age was 56.36 years (Interquartile range 42 to 68 years. 7 (63.63 %) were men and 4 (36.36%) women. Mean time from onset of symptoms to presentation at the day of admission in hospital to death was 7 days [IQR-1-15 days] as mentioned in table given below. Males were n=7(63.6%) and females n=4(36.4%). Co-morbidities were found in all fatal cases Main co-morbid condition was COPD n=5(45.5%) then hypertension n=3(27.3%) and ischemic heart disease n=3(27.3%). During this study period 108 number of our COVID-19 confirmed cases had an outcome that is death or discharge so our case fatality rate was 10.18%. Most common cause of death was Cardiac arrest 7(63.6%) followed by multi organ dysfunction syndrome 2(18.2%) followed by ARDS (Acute respiratory distress syndrome) 2(18.2%).

**Fig 1:**
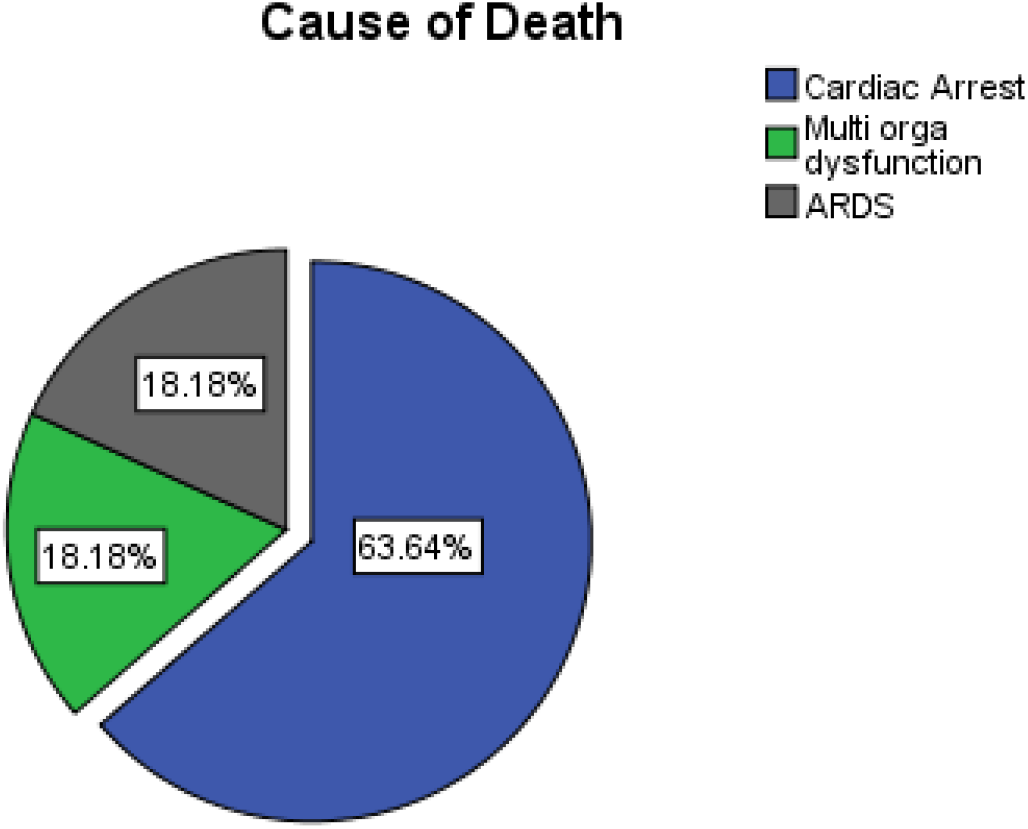
Describing causes of mortality

ARDS, Secondary bacterial infection especially after administration of Tocilizumab, (Tocilizumab is a recombinant humanized anti-interleukin-6 receptor (IL-6R) monoclonal antibody which has a main use in the treatment of rheumatoid arthritis) (10)

Acute kidney injury and acute liver injury were common during hospitalization and lead to a number of ICU admissions. All patients received mechanical ventilation, Corticosteroids (Methyl prednisolone 40 mg intravenous BD) antibiotics including azithromycin. Antibiotics were escalated in 4 cases amid to worsening septic shock. Interestingly 7 cases died suddenly after they had been weaned off from mechanical ventilation and were out of secondary septic shock on acceptable Oxygen supplementation (5-6 liters per minute via simple face masks maintaining their SpO2 at 96 to 97%). They all developed sudden bradycardia then asystole and did not respond to CPR. Cases who could not be anti-coagulated were among those who had survived ARDS and had been weaned off from mechanical ventilation. 4 cases developed severe septic shock and multi-organ failure while 2 cases had refractory ARDS and could not be oxygenated. All cases were given Tocilizumab as they sufficed the criteria of CRS (Cytokines Release syndrome) (11) Biochemical profile analysis revealed that Serum levels of Ferritin, LDH and D-Dimers were higher in non survivors (mean=1370.22).

**Table No: 01.** Characteristics and laboratory results of included fatal cases of COVID-19.

|  |  |  |
| --- | --- | --- |
|  | Mean (max-min) | N (%) |
| **Age** | 56.36 (42-68) |  |
| **Sex** |  |  |
| Male |  | 7(63.6%) |
| Female |  | 4(36.4%) |
| **Number of days** | 7 (1-15) |  |
| **Co-morbidities** |  |  |
| Hypertension |  | 5(45.5%) |
| COPD |  | 3(27.3%) |
| IHD |  | 3(27.3%) |
| **Lab Values** |  |  |
| Serum Ferritin | 1051.41(474-1668) |  |
| *Ferritin >362 ng/ml* |  | 11(100%) |
| Serum LDH | 460.79 (45-683) |  |
| *LDH >220 u/L* |  | 11(100%) |
| D-Dimers | 1370.22 (373-1725) |  |
| *D-Dimers > 500* |  | 10(90.09%) |
| Lymphocyte count | 11.80 (05-68) |  |
| *Lymphocytes < 40%* |  | 10(90.09%) |
| Neutrophils Ratio | 88.87 (86-91) |  |
| *Neutrophils > 80%* |  | 11(100%) |
| ***Tocilizumab*** |  | 11(100%) |
| **Cause of death** |  |  |
| Cardiac Arrest |  | 7(63.6%) |
| Multi organ dysfunction |  | 2(18.2%) |
| ARDS |  | 2(18.2%) |

Following table describe the characteristics and laboratory results of all fatal cases included in this study out of all fatal case 63.6% were male and 36.4% were female the mean age of patients was 56.36 years. The number of days patients remain in hospital were maximum 15 fatal case had co morbidities Hypertension, COPD and IHD but the rate of hypertension was very high 45.5% were with hypertension and 27.3% comorbid cause was COPD and IHD.

The main concern of this case study was the high readings of laboratory values we included in this case some selected parameters of Covid-19 patient’s labs like serum ferritin, LDH, D-Dimers, Lymphocytes, and neutrophils. According to results of this study the average mean of serum Ferritin was 1051, minimum=474, maximum=1668 and these values were higher than normal ferritin in all patients.

Similarly the serum LDH mean value was 460.79, minimum=460, maximum=683 these values were also higher in all cases. It has been noted that serum Ferritin and D-dimers in all the fatal cases stayed above (mean=1051.41) throughout the course of treatment while other markers were observed trending downwards.

## Discussion

11 fatal cases were included. The mean age was 56.36 years (Interquartile range 42 to 68 years. 7 (63.63 %) were men and 4 (36.36%) women. Mean time from onset of symptoms to presentation at the day of admission in hospital to death was 7 days [IQR-1-15 days] as mentioned in table given below. Males were n=7(63.6 %) and females n=4(36.4%). Co-morbidities were found in all fatal cases Main co-morbid condition was COPD n=5(45.5%) then hypertension n=3(27.3%) and ischemic heart disease n=3(27.3%). During this study period 108 number of our COVID-19 confirmed cases had an outcome that is death or discharge so our case fatality rate was 10.18% Most common cause of death was Cardiac arrest 7(63.6%) followed by multi organ dysfunction syndrome 2(18.2%) followed by ARDS (Acute respiratory distress syndrome) 2(18.2%).

A study conducted in china found that almost half patients with COVID-19 wee over the age of fifty years, which men were more likely to be infected than women the death rate in males is above that in females. In patients who develop SARS, advanced age was an independent predictor for an adverse outcome, but sex was not, the foremost common comorbidities of the patients with COVID-19 in our cohort are hypertension and diabetes, which is analogous to findings of previous studies, however, the foremost common comorbidities within the patients with SARS were diabetes (16 [11%]) and cardiac disease (12 [8%])(12).

A recent study by Weiss and colleagues of 191 patients, of whom 54 died, found that older age, high sequential organ failure assessment score, and D-dimer >1 μg/ml could assist within the early identification of patients who may have a poorer prognosis. In our study, the median age of nonsurvivors was 65.8 years, which is analogous to the median age reported in non-survivors, but more than that of the survivors (52 yr), reported within the previous study. A study found that the median (SD) level of D-dimer in non-survivors was 5.159 (4.679) μg/ml (range, 0.27–26 μg/ml), and 70.6% of our patients had a D-dimer >1 μg/ml, which was also according to the previous study(13). In our study It has been noted that serum Ferritin and D-dimers in all the fatal cases stayed above (mean=1051.41) throughout the course of treatment while other markers were observed trending downwards.

## Data Availability

All data is available with the IRB&EC of Hospital

